# Autoimmune and Inflammatory Mechanisms in Cervical Dystonia

**DOI:** 10.1101/2020.09.03.20187815

**Authors:** Gamze Kilic-Berkmen, Laura Scorr, Ashok R. Dinasarapu, Lucas McKay, Ami Rosen, Pritha Bagchi, John Hanfelt, Andrew McKeon, H. A. Jinnah

## Abstract

There are many causes for cervical dystonia (CD), although most cases are idiopathic and a cause cannot be identified. The observation that 10-15% of cases have an affected family member has pointed to genetic causes, but known genes account for only a small fraction of all cases. The current manuscript describes a series of studies focusing on potential autoimmune or inflammatory mechanisms in CD. First, a case-control survey for 32 autoimmune diseases in 271 subjects with CD confirmed prior anecdotal observations that CD is associated with thyroid disease, which often results from autoimmune mechanisms. Second, unbiased proteomic methods involving a total of 20 subjects with CD, with or without associated thyroid disease, pointed towards a series of overlapping mechanisms relating to the immune system. Third, a multiplex immunoassay focusing on 37 markers associated with neuroinflammation applied to a total of 20 subjects with CD with or without thyroid disease and 20 controls pointed to abnormalities in several specific measures of the immune system. Finally, a broad screening test for neuronal antibodies in a total of 58 subjects with CD did not disclose any specific antibodies. Altogether, the association of CD with thyroid disease and blood-based immune measures point to abnormalities in cell-mediated immunity that may play a pathogenic role for a subgroup of subjects with CD.

## Introduction

Cervical dystonia (CD) is characterized by excessive involuntary contraction of neck muscles leading to abnormal postures and movements of the head, often with neck pain^21^. Etiologies are heterogeneous and include focal or degenerative lesions of the nervous system, exposure to drugs or medications, infections, and others. However, most cases are idiopathic and a cause cannot be found. Approximately 10-15% of cases have an affected family member. This observation points to inherited mechanisms, but known genes explain only a small fraction of cases^26^.

In view of these observations, there is a need to consider non-genetic mechanisms in CD. In fact, multiple anecdotal reports link CD and related focal dystonias with immune mechanisms^4, 5^. Several reports have described autoimmune thyroid disease in individuals with CD or related focal dystonias ^3, 25, 28, 30, 33, 34^. Multiple uncontrolled case series have suggested autoimmune thyroid disease to be more frequent in CD and related conditions than expected^6, 9, 11, 19, 20^. CD has also been linked with other autoimmune disorders including multiple sclerosis^1, 7, 29, 38, 39, 41^, systemic lupus erythematosus^37^, Sjogren syndrome^27, 43^, and myasthenia gravis ^14,15, 18^. CD and other related focal dystonias have been reported as a delayed consequence of various infections, a phenomenon often caused by immunological cross-reactivity between the infectious agent and host targets.8, 12, 16, 17, 23, 24, 32, 36

These observations raise the possibility that immune mechanisms may play a pathogenic role for a subgroup of CD. Few studies have methodically explored this possibility. One study of 11 subjects with CD described abnormalities in the proportions of helper and suppressor T-lymphocytes^31^. Another study of 75 CD subjects identified a link with the HLA DQB1 allele^10^. A few subjects with CD were reported to respond to corticosteroid immunosuppressants^12, 27^. A study of 65 subjects with a variety of adult-onset dystonias concluded that anti-basal ganglia antibodies were not common, but may be relevant for a subgroup^13^.

In the current studies, a potential relationship between CD and immune mechanisms was explored via several independent methods that included a methodical survey of autoimmune disorders in CD, unbiased shotgun proteomics of blood samples, serological studies for immunological markers, and broad screens for neuronal antibodies.

## Methods

### Subjects

The study was approved by the Emory University IRB and all subjects gave informed consent to participate. All subjects were at least 18 years of age. Different cohorts of subjects were used for the studies below. To be included in the idiopathic CD group, the participant had to have a diagnosis of CD. In addition to CD, they could have mild dystonia in other regions with segmental or multifocal patterns. People with other neurologic diagnoses were excluded from this group. In addition, people with CD secondary to known causes such as focal brain lesions, neuroleptic exposure, or parkinsonism. Cases with known genetic causes for their CD were also excluded from the study.

For the cross-sectional survey study, the control group included disease controls presenting to the movement disorders clinic for problems other than dystonia (e.g., Parkinson disease or parkinsonism, tremor, chorea, ataxia, and various neurogenetic syndromes). For all blood-based studies, the control group included only neurologically normal individuals, to minimize sample variance. Control subjects with known autoimmune disorders (e.g. multiple sclerosis) were excluded from the blood-based studies.

### Autoimmunity survey

With input from two experienced neuro-immunologists, a questionnaire was designed to survey the most common autoimmune disorders, along with some rarer subtypes relevant to other neurological disorders. Most subjects and controls for this study were recruited from Movement Disorders Clinics, or from patient support group conferences. They were first asked if they would participate in a 5-minute survey. For all who agreed to be interviewed, they were next asked if they had any known autoimmune disorders. Following this initial open question, subjects were asked to indicate if they or any first-degree relatives had any of 32 specific autoimmune disorders from the list (Table 1). Finally, they were asked if they or their relatives had any other autoimmune disorders not on the list.

**Table 1.** Autoimmune disorders in CD

| Condition | Controls (n=254) | CD (n=271) |
| --- | --- | --- |
| Neurological |  |  |
| Multiple sclerosis | 0 | 2 (1%) |
| Myasthenia gravis | 0 | 3 (1%) |
| Guillain-Barre syndrome | 0 | 0 |
| CIDP | 0 | 0 |
| Polymyositis | 0 | 0 |
| Dermatomyositis | 0 | 0 |
| Endocrine |  |  |
| Hypothyroidism (under-active) | 15 (6%) | 46 (17%) |
| Hyperthyroidism (over-active) | 6 (2%) | 8 (3%) |
| Hashimoto thyroiditis | 2 (1%) | 6 (2%) |
| Grave's disease | 0 | 1 (0.4%) |
| Diabetes mellitus, Type 1 | 0 | 2 (1%) |
| Addison's disease | 0 | 0 |
| Adrenitis (autoimmune) | 0 | 0 |
| Skin |  |  |
| Vitiligo | 0 | 2 (1%) |
| Scleroderma | 0 | 0 |
| Pemphigus | 0 | 0 |
| Psoriasis | 12 (15%) | 9 (3%) |
| Gastrointestinal |  |  |
| Crohn's disease | 2 (1%) | 0 |
| Ulcerative colitis | 3 (1%) | 5 (2%) |
| B12 deficiency | 16 (6%) | 17 (6%) |
| Bones & joints |  |  |
| Rheumatoid arthritis | 5 (2%) | 9 (3%) |
| Reactive arthritis (Reiter's syndrome) | 0 | 0 |
| Ankylosing spondylitis | 2 (1%) | 0 |
| Systemic |  |  |
| Lupus (SLE) | 0 | 0 |
| Sjogren's syndrome | 0 | 1 (0.4%) |
| Hematologic |  |  |
| Hemolytic anemia | 2 (1%) | 0 |
| Aplastic anemia | 0 | 1 (0.4%) |
| Pernicious anemia | 0 | 2 (1%) |
| Antiphospholipid syndrome | 0 | 0 |
| Thrombocytopenia | 0 | 0 |
| TTP | 0 | 0 |
| ITP | 0 | 0 |
| Other autoimmune disorders | 6 (2%) | 13 (5%) |
| No autoimmune disorders | 181 | 140 |

To avoid biasing answers, the interviewer was instructed to read only the name of the disorder listed, without giving any qualifying explanations. Because many subjects did not know the cause of their thyroid disease, and because autoimmune hyperthyroidism may evolve to hypothyroidism, all thyroid disorders were combined into a single category. In cases where the subject reported the same disorder in two different ways (e.g. Hashimoto thyroiditis plus hypothyroidism or Grave’s disease plus hyperthyroidism), only the more specific diagnosis was retained. Non-autoimmune thyroid conditions caused by surgery, trauma, tumors were excluded. The questionnaire divided diabetes mellitus into types 1 and 2 to ensure recording of the correct form, but type 2 was not included in the final statistical analyses focusing on autoimmune disorders because it is not usually an autoimmune disorder. A preliminary study with power analysis indicated the need to interview at least 250 subjects with CD and 250 controls for a statistically meaningful result.

Descriptive statistical analysis was performed for demographic characteristics including age and sex. A Chi-square test was used to evaluate whether autoimmune disorders were more common in CD compared to controls. All tests were 2-sided and p-values < 0.05 were regarded as significant.

### Unbiased shotgun proteomics

Unbiased proteomic methods were applied to plasma samples collected from subjects with CD or normal controls. When subjects with idiopathic CD are studied as a group, etiological heterogeneity hampers efforts to identify subgroups that may have specific causes. Therefore, CD subjects with coincidental thyroid disease (CD+T) were evaluated separately from those without thyroid disease (CD-T) to enrich for a potential subgroup with autoimmune mechanisms. To reduce sample variance, the study was limited to females with minimal comorbidities and similar ages (CD-T, 66.3±10.6 years; CD+T, 65.6±8.7 years, controls, 57.0±12.5 years). To limit any influence of CD treatments, all blood samples were collected approximately 3 months following prior Botulinum toxin treatments.

Within 4 hrs of blood draw, plasma was separated and frozen in 0.2 mL aliquots and stored at −80°C. Further processing was done by the Emory Integrated Proteomics Core. In brief, samples were first processed to remove 14 of the most abundant proteins, a procedure that permits detection of a larger number of lower abundance proteins. Samples were then subjected to partial in-solution trypsin digestion and analyzed by liquid chromatography with tandem mass spectrometry (LC-MS/MS) on an Orbitrap Fusion mass spectrometer.

Peptide-specific ion intensities were extracted to derive relative protein abundance using MaxQuant (v1.6.10.43) label-free quantification. All raw files were processed together in a single run. Database searches were performed using the Andromeda search engine (a peptide search engine based on probabilistic scoring) with the UniProt-SwissProt human canonical database as a reference and a contaminants database of common laboratory contaminants. MaxQuant employs the MaxLFQ algorithm for label-free quantitation (LFQ). Quantification was performed using razor and unique peptides, including those modified by acetylation (protein N-terminal), oxidation or deamidation. Protein expression (quantification) was determined from LFQ intensities calculated by MaxQuant. LFQ data processing was further done with Perseus (v1.6.10.50). Known common contaminants and LFQ values with very low expression were filtered out. Samples with > 50% missing values were excluded. Any remaining missing LFQ values were imputed and LFQ intensities were log2-transformed to reduce the effect of outliers. A Protein Group consists of identified peptides that are shared across multiple proteins. For downstream analyses we considered only the main protein of a Protein Group. Log ratios were calculated as the difference in average log2 LFQ intensity values between groups.

Protein expression levels were assessed with linear regression models, with CD as a dichotomous variable. Additional linear models were calculated incorporating an interaction term coding for an additive effect of thyroid disorder. Because this was an exploratory study, initial statistical comparisons focused on two-tailed Student’s t-tests with p< 0.05 regarded as significant. Shotgun proteomic studies often do not employ typical methods such as the False Discovery Rate (FDR) to correct for multiple comparisons, because they are considered too conservative^35^. However, p-values corrected for FDR< 0.10 are presented for reference. All proteins with a nominal p< 0.05 were subjected to pathway analyses to search for common biological pathways. Network analyses were conducted using STRING, a database of known and predicted protein-protein interactions (https://string-db.org). The interactions include direct (physical) and indirect (functional) associations. These interactions were determined from interactions aggregated from other (primary) databases, from knowledge transfer between organisms, and computational predictions.

Variations in protein abundance were assessed with linear regression models. Main models included CD as a dichotomous variable. No formal adjustment for multiple comparisons was performed in primary analyses. P-values adjusted for False Discovery Rate (FDR) are presented for reference. Additional linear models were calculated incorporating an interaction term coding for the additive effect of thyroid disorder in addition to that of CD.

### Plasma multiplex immunoassays

A total of 37 plasma markers associated with neuroinflammatory markers and related immunological processes were quantified using a multiplex electrochemiluminescence-based immunoassay (MesoScale Discover, Gaithersburg, MD). To reduce sample variance, this study was again limited to females with CD (with or without thyroid disease) and normal controls with minimal comorbidities and similar ages (CDT, 66.3±10.6 years; CD+T, 67.5±8.3 years; controls, 65.2±10.9 years.

The assay evaluated 37 markers of inflammation including pro- and anti-inflammatory cytokines and chemokines (Table 2). Following statistical evaluation to confirm the absence of outliers, descriptive statistics for each analyte were calculated in a stratified approach: 1) stratification by CD vs control, and, 2) stratification by CD+T and CD-T vs. control. Variations in the expression of each analyte were assessed with linear regression models, with CD as a dichotomous variable. Additional linear models were calculated incorporating an interaction term coding for the additive effect of thyroid disorder in addition to that of CD. No formal adjustment for multiple comparisons was performed, but p-values adjusted for FDR are presented for reference.

**Table 2.** Multiplex immunoassay

|  | CD-T (n=10) | CD+T (n=10) | Controls (n=20) |
| --- | --- | --- | --- |
| **Pro-inflammatory measures** |  |  |  |
| IFN-γ | 4.19 (3.61) | 15.20 (30.0) | 3.10 (2.57) |
| IL-1β | 0.06 (0.07) | 0.17 (0.23) | 0.07 (0.06) |
| IL-2 | 0.27 (0.21) | 0.61 (0.49) | 0.26 (0.19) |
| IL-4 | 0.01 (0.01) | 0.02 (0.02) | 0.01 (0.01) |
| IL-6 | 1.10 (1.11) | 0.93 (0.48) | 1.02 (0.60) |
| IL-8 | 6.29 (9.81) | 7.49 (11.5) | 3.68 (2.36) |
| IL-10 | 0.39 (0.30) | 0.91 (0.89) | 0.33 (0.36) |
| IL-13 | 0.78 (0.55) | 1.96 (1.79) | 1.34 (1.38) |
| TNF-α | 2.77 (0.71) | 3.76 (1.23) | 3.46 (1.02) |
| **Cytokines** |  |  |  |
| IL-1α | 5.83 (5.68) | 11.30 (19.20) | 3.51 (3.05) |
| IL-5 | 0.67 (0.232) | 0.82 (0.39) | 0.52 (0.37) |
| IL-7 | 2.56 (2.63) | 2.66 (2.00) | 2.19 (1.02) |
| IL-12/IL-23p40 | 169 (87.70) | 228 (83.0) | 231 (125.0) |
| IL-15 | 2.64 (0.39) | 2.81 (0.60) | 2.67 (0.47) |
| IL-16 | 190 (32.2) | 199 (71.0) | 202 (67.3) |
| IL-17A | 1.92 (1.06) | 2.43 (0.64) | 2.57 (2.29) |
| TNF-β | 0.19 (0.10) | 0.28 (0.14) | 0.22 (0.09) |
| VEGF-A |  |  |  |
| **Chemokines** |  |  |  |
| Eotaxin | 149 (62.7) | 129 (58.8) | 136 (53.0) |
| MIP-1β | 22.5 (6.33) | 46.4 (75.2) | 38.5 (35.1) |
| Eotaxin-3 | 90.1 (15.9) | 202 (428) | 80.2 (23.9) |
| TARC | 23.9 (34.6) | 15.7 (14.1) | 9.83 (6.41) |
| IP-10 | 263 (142) | 378 (337) | 308 (216) |
| MIP-1α | 15.9 (3.01) | 16.1 (4.23) | 18.0 (4.68) |
| MCP-1 | 16.5 (3.79) | 25.2 (31.8) | 18.5 (3.48) |
| MDC | 313 (82.9) | 276 (70.1) | 320 (91.2) |
| MCP-4 | 223 (164) | 183 (53.9) | 174 (52.2) |
| **Angiogenesis** |  |  |  |
| VEGF-C | 46.9 (16.7) | 46.3 (50.9) | 52.4 (35.2) |
| VEGF-D | 877 (236) | 789 (119) | 707 (290) |
| Tie-2 | 3390 (506) | 3150 (884) | 3560 (715) |
| Flt-1 | 62.8 (15.8) | 60.8 (11.6) | 56.9 (15) |
| PIGF | 6.40 (1.69) | 6.08 (1.40) | 5.07 (1.11) |
| bFGF | 18.1 (21.4) | 17.2 (16.3) | 26.9 (32.5) |
| **Vascular injury** |  |  |  |
| SAA | 6.55x10<sup>6</sup><br>(8.04x10<sup>6</sup>) | 6.51x10<sup>6</sup><br>(4.60x10<sup>6</sup>) | 8.06x10<sup>7</sup><br>(1.24x10<sup>7</sup>) |
| CRP | 3.99x10<sup>6</sup><br>(2.92x10<sup>6</sup>) | 1.90x10<sup>6</sup><br>(1.35x10<sup>6</sup>) | 6.53x10<sup>6</sup><br>(6.96x10<sup>6</sup>) |
| VCAM-1 | 4.07x10<sup>5</sup><br>(1.37x10<sup>5</sup>) | 3.98x10<sup>5</sup><br>(6.52x10<sup>4</sup>) | 4.35x10<sup>5</sup><br>(1.17x10<sup>5</sup>) |
| ICAM-1 | 4.56x10<sup>5</sup><br>(1.30x10<sup>5</sup>) | 4.08x10<sup>5</sup><br>(6.20x10<sup>4</sup>) | 4.70x10<sup>5</sup><br>(1.96x10<sup>5</sup>) |

NB: Values for analytes reflect concentrations calculated from electroluminescence assay in pg/ml

### Serum auto-antibodies

Serum samples were evaluated for potential auto-antibodies using a standardized indirect immunofluorescence assay for IgG binding to neuronal and glial cells in mouse brain, kidney, and gut^22^. The assay included antineuronal nuclear antibody type 1 (ANNA-1 or anti-Hu), antineuronal nuclear antibody type 2 (ANNA-2 or anti-Ri), antineuronal antinuclear antibody type 3 (ANNA-3), and anti-glial and/or neuronal nuclear antibody type 1 (AGNA-1). The assay included antibodies that bind to neuronal cytoplasm (PCA types 1 and 2), collapsin-response mediator protein 5 (CRMP-5), and amphiphysin. Finally, it included antibodies targeting neuronal cell surface receptors delta-notch epidermal-like growth factor– related receptor (PCA-Tr) and metabotropic glutamate receptor 1 (mGluR1). The reference values for titers for all antibodies were < 1:240.

Serum samples also were tested by radioimmunoprecipitation assays for antibodies reactive with neural cation channel complexes including neuronal voltage-gated calcium channels (P/Q type and N type), voltage-gated potassium channel (VGKC) complexes, and nicotinic acetylcholine receptors (AChR) of skeletal muscle type [α_1_ subunit] and neuronal ganglionic type [α_3_ subunit]) and glutamic acid decarboxylase 65-KDa isoform (GAD65). Reference values for all antibodies were no greater than 0.02 nmol/L (with the exception of the N-type calcium channel antibody, which was ≤0.03 nmol/L). Serum samples also were tested for striational antibodies of the skeletal muscles (by enzyme-linked immunosorbent assay; the reference value was < 120 nmol/L) and CRMP-5 IgG (by recombinant Western blot assay).

## Results

### Autoimmunity survey

Because autoimmune disorders often co-occur^2^, a survey was conducted to determine if autoimmune disorders were more common in idiopathic CD than in controls. Among the 32 autoimmune diseases considered (Table 1), thyroid disease was reported by 23% of subjects with CD (n = 61 of 271) compared to 9% of controls (n = 23 of 254). This difference was statistically significant (p = 0.0001). Subjects with CD also reported high frequencies of thyroid diseases in their first-degree family members, but the numbers of cases were too small for statistical comparisons. Other autoimmune diseases were reported with similar frequencies in CD and controls. These results confirm prior anecdotal reports that thyroid disease occurs with significantly greater frequency than expected in CD compared to controls.

### Unbiased plasma proteomics

Plasma samples were evaluated by unbiased shotgun proteomics to explore the possibility of underlying autoimmune or inflammatory mechanisms. Because of etiological heterogeneity in CD, normal controls (N = 10) were compared to CD subjects, with (CD+T, N = 10) and without (CD-T, N = 10) coincidental thyroid disease, to enrich for a CD subgroup with potential immunological mechanisms. A total of 536 protein groups were identified across the entire cohort of 30 subjects. After filtering potential contaminants and exluding protein groups with > 50% missing values, 213 protein groups remained for final quantification.

The data were evaluated by linear regression, with comparison groups being normal controls, or CD with or without thyroid disease. This analysis revealed significant differences between CD (n = 20 combined) and controls (n = 10), with no additional influence of thyroid disease. Comparisons of all CD combined (n = 20) and controls (n = 10) revealed significant differences for 25 proteins at p< 0.05, and 3 remained significantly different at FDR< 0.10. Results for all group comparisons are shown in Figure 1, along with a volcano plot and boxplots of protein abundance for the 3 most significantly affected proteins (DBH, F13A1, and HGFAC).

**Figure 1.**
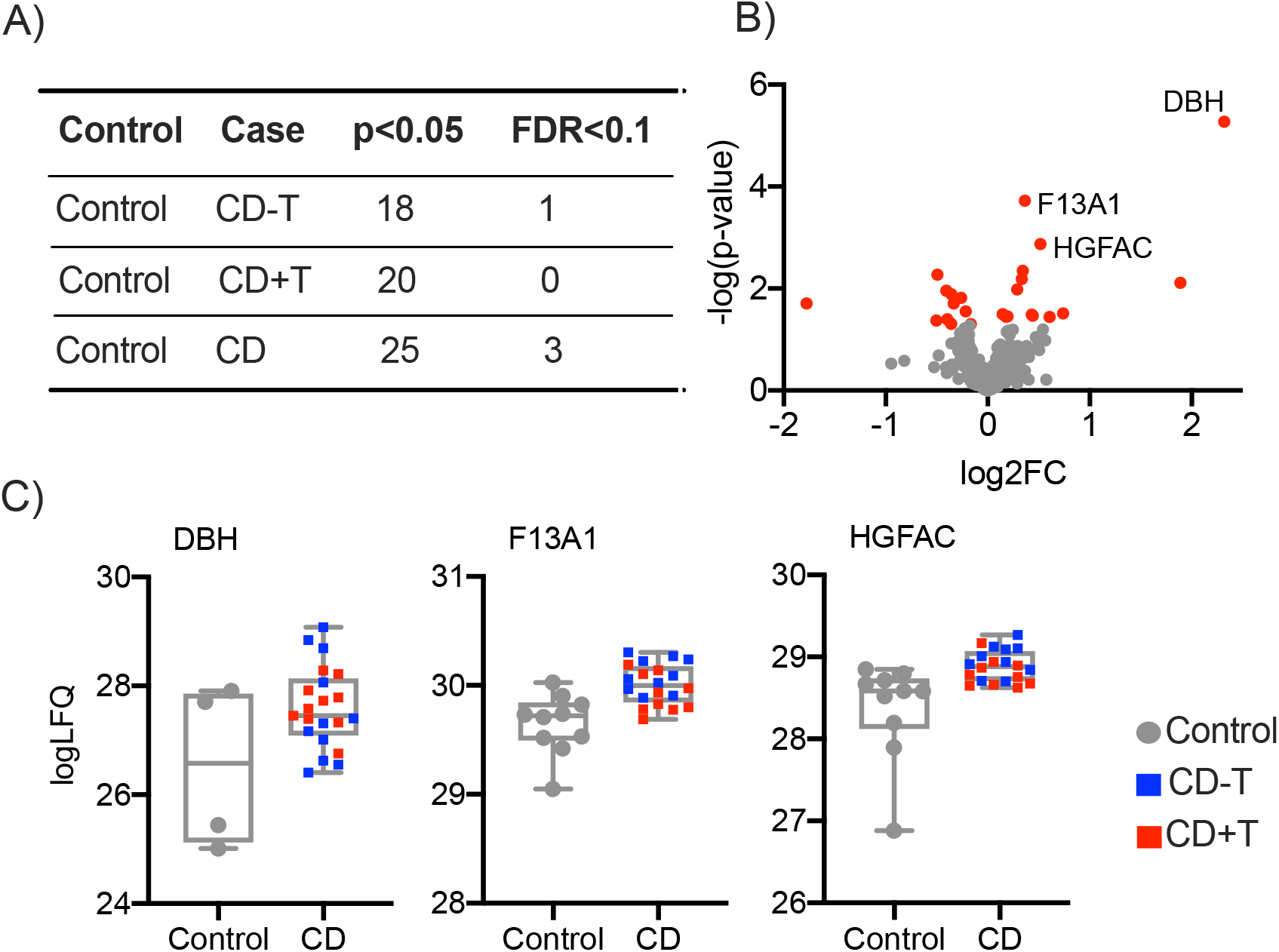
Proteomics in cervical dystonia. Panel A shows the number of differentially expressed proteins (DEPs) between CON and CD-T, CON and CD+T and, CON and CD. CD represents all CD individuals including CD-T and CD+T. Panel B shows the volcano plot with DEPs (red color) of CON and CD comparison. The nominal p-value is shown on the Y-axis and the fold change (FC) is shown on the X-axis. Panel C illustrates protein abundance in box plots of selected DEPs of CON and CD comparison. Abbreviations: CRP: C-reactive protein, DBH: Dopamine beta-hydroxylase, TLN1: Talin-1, FETUB: Fetuin-B, F13A1: Coagulation factor XIII A chain, HGFAC: Hepatocyte growth factor activator, SAA: Serum amyloid A-4 protein, CD44: CD44 antigen, LBP: Lipopolysaccharide-binding protein, AGT: Angiotensin-1.

Considering the 25 proteins with p< 0.05, the top most significant STRING biological pathways identified when comparing CON vs CD were acute inflammatory response, acute phase response, blood coagulation-fibrin clot formation, protein activation cascade, and response to stress (all pathways FDR< 0.001).

In summary, the proteomics study revealed 25 proteins to be abnormal at p< 0.05, and 3 survived stringent testing for multiple comparisons at FDR< 0.10. Although 5 different pathways were identified from the STRING database, the proteins involved overlapped, and most were involved in immune responses. The three proteins with largest magnitude changes (DBH, TLN1, CRP) all are involved in immune responses.

### Multiplex immunoassays

Unbiased proteomic methods are valuable because they provide a broad overview of any potential abnormalities, but they risk overlooking biologically relevant changes because of statistical methods commonly used to address multiple comparisons^35^, and many low abundance proteins may not be detected. Therefore, plasma samples also were evaluated using a more specific multiplex immunoassay targeting 37 analytes associated with immune and inflammatory mechanisms (Table 2).

The serum concentrations for 37 analytes were compared first in subjects with CD (n = 20) versus controls (n = 20) regardless of thyroid status. This analysis revealed the CD group to have significantly decreased CRP (p = 0.03) and increased IL-5 (p = 0.05), and trends towards decreased IL-10 (p = 0.08), increased IL-2 (p = 0.09), and increased PIGF (p = 0.09) (Table 2 and Figure 2). The CD cohort continued to demonstrate a predominantly pro-inflammatory profile when accounting for presence of thyroid disease, confirming the effects were not mediated solely by presence of coincidental thyroid disease, with significantly increased PIGF (p = 0.026), trends towards increased TARC (p = 0.063) and increased VEGF-D (0.081). Supporting the hypothesis that subjects with CD and autoimmune hypothyroid (CD+T) disease may represent a subgroup with a unique biological substrate, a number of biomarkers differed significantly in the interaction of CD and thyroid disease such that cases of CD+T included significantly increased IL-4 (p = 0.011), increased IL-2 (0.015), increased IL-10 (0.036), increased TNF-alpha (0.036). There were also trends towards increased IL-1beta (0.072) and increased TNF-beta (0.082).

**Figure 2.**
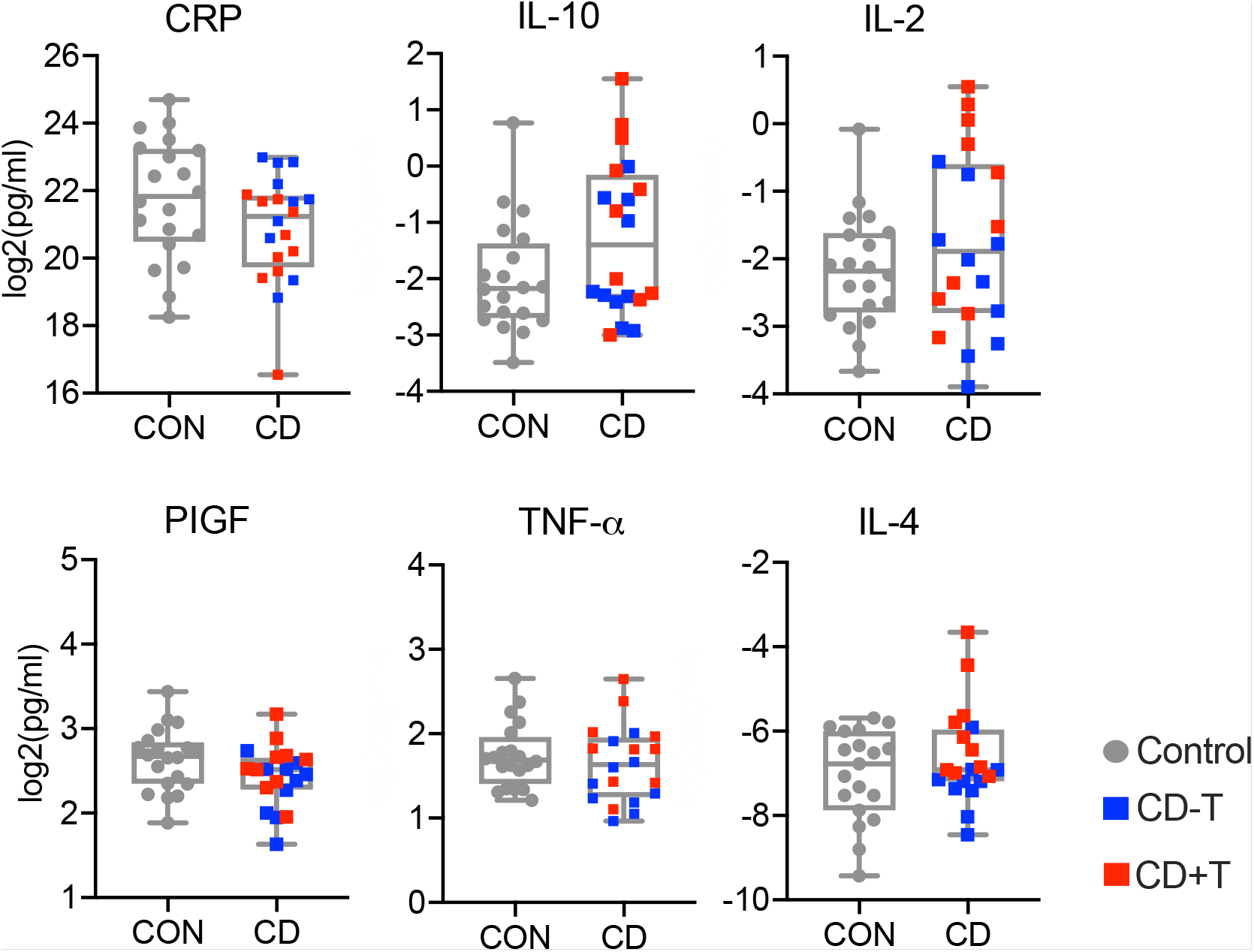
Serum Immune Profile. Pro- and anti-inflammatory chemokines and cytokines differed significantly among CD cases and controls. Supporting the hypothesis that subjects with CD and autoimmune hypothyroid (CD+T) disease may represent a subgroup with a unique biological substrate, a number of biomarkers differed significantly in the interaction of CD and thyroid disease such that in cases of CD+T there was significantly increased IL-4 (p = 0.011), increased IL-2 (0.015), increased IL-10 (0.036), increased TNF-alpha (0.036). There were also trends towards increased IL-1beta (0.072) and increased TNF-beta (0.082).

### Serum autoantibodies

Finally, serum samples were evaluated for autoantibodies using a tissue-based assay. The subjects in this cohort included 30 CON, 30 CD-T, and 28 CD+T. For this analysis, 30 cases with autoimmune thyroid disease but without CD (T-CD) were also available. In the initial screen, 2 of the 30 cases with CD-T had low titers of neuronal antibodies. The pattern was not specific for any known neuronal antibodies, and was not replicated in a second serum sample from each. None of the other cases had any detectable neuronal antibodies.

GAD65 antibodies were found for 9/30 CON, 11/28 cases of CD-T, 6/30 cases of CD+T, and 9/30 T-CD. For all of these cases, GAD65 antibody titers were always < 5 nmol/L. These antibodies are common in the general population (approximately 8% in normal individuals) and only titers > 20nmol/L are considered significant. Gastric anti-parietal cell antibodies were found in low titers for 5/30 cases of CD+T. They were not seen in any of the 28 cases of CD-T, nor in any CON. Overall, these studies do not provide evidence for pathological titers of any of the common autoimmune antibodies in CD.

## Discussion

CD is a heterogeneous disorder with many potential causes, although the vast majority of cases are idiopathic, even after exhaustive evaluation. When subjects with idiopathic CD are studied as a group, etiological heterogeneity hampers efforts to identify subgroups with potentially distinct biological causes. Many studies have explored genetic causes, because 10-15% of cases have an affected family member. These genetic studies are successful in identifying a cause in only a small fraction of cases^26^. Therefore, additional potential causes should be considered. The current manuscript describes a series of methods exploring immune mechanisms. To identify a potential CD subgroup where this etiology might be relevant, a strategy was employed that focused on the relationship between CD and thyroid disease.

The autoimmunity survey demonstrated that thyroid disease was significantly more frequent in CD compared with other neurological controls, a finding that confirms prior anecdotal observations. Although the survey data could not specify a cause for thyroid disease in all cases, most adult-onset cases of primary hypothyroidism arise from autoimmune mechanisms^2^. The unbiased proteomics study pointed to potential abnormalities of the immune system, and the multiplex immunoassay identified abnormalities among several specific immune markers. However, no specific neuronal antibodies were found. These results provide useful guidance for further studies of potential immune mechanisms in CD.

The failure to find any relevant neuronal antibodies in CD is consistent with a prior report that focused on anti-basal ganglia antibodies across many types of dystonia^13^. The current studies extend these prior findings to include antibodies that may target other brain regions, using a more homogeneous population of CD cases, including a sub-population of CD with coexisting thyroid disease. Although the numbers of cases in both antibody screens were relatively small, the results suggest larger screens using serum would not be fruitful, although studies of neuronal antibodies in cerebrospinal fluid may be informative. This observation suggests that a more fruitful approach may involve studies of other components of the immune system such as immune cell repertoires, cytokines, or chemokines. Additional attention to T-cells may be warranted, because the majority of cases with autoimmune thyroid disease are caused by auto-reactive T-lymphocytes rather than antibodies^2, 40^. In fact, several of the differentially expressed immune markers identified in the multiplex assay are produced by T-cells including IL-2, IL-4, IL-5, and IL-10. Together, these observations suggest that further attention should be directed towards cell-based immune mechanisms, and particularly T-cells.

Shotgun proteomics are attractive because they have the potential to disclose numerous biological mechanisms, unbiased by preconceived hypotheses. However, their power to detect abnormalities is hampered by the statistical procedures commonly used address multiple comparisons^35^, as well as a known bias for detection of proteins expressed at relatively high levels. Although the current findings should be considered preliminary because of the small numbers of cases evaluated, they consistently pointed toward abnormalities in overlapping pathways involving the immune system that should be further explored in more targeted studies. Such studies may require large numbers of subjects. A more targeted approach, such as multiplex immunoassays focusing on specific mechanisms, may be more efficient. In fact, this method pointed to more specific markers of the immune system that were not identified by shotgun proteomics. Only 4 of the proteins in the multiplex panel (CRP, ICAM1, SAA[1-4] and VCAM1) were also measured in the proteomics study, most likely because of low abundance or technical issues with resolution using LC-MS. One of these 4 proteins was significantly altered in both assays (SAA[1-4]).

The greatest limitation of the current studies is the relatively small numbers of cases studied. In order to identify a subgroup where immunological mechanisms are relevant, it may be necessary to study a much larger group. CD is not a common disorder, so collecting large numbers of cases is a challenge. Finding subjects that have both CD and thyroid disease is even more challenging. In this situation, a multicenter collaborative effort may be required. Another weakness of the current studies relates to the use of blood samples to elucidate mechanisms for a brain disorder. Studies of cerebrospinal fluid or brain tissue itself may be required, although large collections of samples from subjects with CD are not currently available. A third weakness relates to observations that the diagnosis of CD is often delayed for many years^42^. The collection of blood samples many years after onset of CD will miss any transient immunological processes occurring early when it first began. A fourth weakness is that prior treatment with Botulinum toxin may influence immunological measures, even though samples were collected 3 months after any treatment. Finally, at first glance, the results suggestive of immune mechanisms in CD seem to conflict with other evidence for genetic causes. On the other hand, immune mechanisms and autoimmunity are both inherited^2^, potentially explaining the apparently low penetrance in many families. In addition, some genetic disorders are well known to trigger secondary immune responses.

Although immunological mechanisms may account for only a small subgroup of CD, identifying this subgroup is clinically important because immune-based therapies may be useful. The current results do not support trials of such therapies at present until such a subgroup and the immune mechanisms can be delineated more precisely. Neurological disorders resulting from antibody-based mechanisms (e.g. Guillan-Barre syndrome) respond to plasmapheresis or high-dose intravenous immunoglobulin, while disorders resulting from autoreactive lymphocytes (e.g. multiple sclerosis) require corticosteroids or more aggressive immunosuppressants. Both types of treatments are associated with substantial potential toxicity.

## Data Availability

Data utilized for these manuscripts is available upon request.

## Acknowledgements

This work was supported in part by grants to the Dystonia Coalition (U54 NS065701, U54 TR001456, and U54NS116025), a consortium of the Rare Diseases Clinical Research Network (RDCRN) that is supported by the Office of Rare Diseases Research (ORDR) at the National Center for Advancing Clinical and Translational Studies (NCATS) in collaboration with the National Institute for Neurological Diseases and Stroke (NINDS).

